# Quantifying Associations Between Child Health and Neighborhood Social Vulnerability: Does the Choice of Index Matter?

**DOI:** 10.1101/2023.06.20.23291679

**Authors:** Anna Zolotor, Ro W. Huang, Nrupen A. Bhavsar, Rushina Cholera

## Abstract

**Importance:** Policymakers have increasingly utilized place-based social disadvantage indices to quantify the impacts of place on health and inform equitable resource allocation. Indices vary in design, content, and purpose but are often used interchangeably, potentially resulting in differential assignments of relative disadvantage depending on index choice.

**Objective:** To compare associations between three commonly used disadvantage indices (Social Vulnerability Index (SVI), Area Deprivation Index (ADI), and Child Opportunity Index (COI)) and two epidemiologically distinct child health outcomes—infant well-child check (WCC) attendance and adolescent obesity.

**Design:** Cross-sectional analysis of Duke University Health System electronic health record (EHR) data from January 2014 to December 2019.

**Participants:** Children ≤18 years of age with outpatient encounters between January 2014 and December 2019, and who were Durham County residents were eligible. WCC attendance was assessed for infants ages 0-15 months; obesity was assessed for children ages 11-17 years.

**Exposures:** 2014 Social Vulnerability Index (SVI), 2015 Area Deprivation Index (ADI), and 2015 Child Opportunity Index (COI) 2.0.

**Main Outcomes:** 1) Infant WCC attendance: attending less than the minimum recommended six WCCs in the first 15 months of life, and 2) Adolescent obesity: BMI ≥ the 95th percentile at both the most recent encounter and an encounter within the prior 9-36 months.

**Results:** Of 10175 patients in the WCC cohort, 20% (n = 2073) had less than six WCCs. Of 14961 patients in the obesity cohort, 20% (n = 2933) had obesity. All three indices were associated with both WCCs (OR for SVI 1.10, 95% CI 1.08-1.12; OR for ADI 1.10, 95% CI 1.08-1.12; OR for COI 1.12, 95% CI 1.10-1.14) and obesity (OR for SVI 1.05, 95% CI 1.04-1.08; OR for ADI 1.08, 95% CI 1.06-1.10; OR for COI 1.07, 95% CI 1.05-1.08).

**Conclusions and relevance:** Higher social disadvantage as defined by all three indices was similarly associated with both adolescent obesity and decreased infant WCC attendance. While the COI incorporates a broader set of child-specific variables, the SVI and ADI may often be just as suitable for pediatric research. Users should consider population and outcome characteristics when selecting an index.

## Introduction

Health-related social needs (HRSNs), such as access to nutritious food or safe and stable housing, shape health outcomes in childhood and across the lifespan.^1^ HRSNs affect children differently than adults given the importance of early life experiences on developmental trajectory and lifelong health,^2^ increased susceptibility to environmental exposures,^3^ and dependence on caregivers. HRSNs are not uniformly distributed across populations due to multi-level structural oppression that leads to inequities in resources, power, and ultimately health.^4^ Area-level variables, such as median income of a census tract, have been frequently used to provide insight into the distribution of HRSNs within neighborhoods.^5^

In the last decade, however, researchers and policymakers have increasingly utilized social disadvantage indices to assess the impact of neighborhood environment on health and to inform resource allocation to communities with greater need.^6,7^ By combining multiple population-level demographic and economic variables, social disadvantage indices provide a more robust picture of place-based social vulnerability than single socioeconomic measures.

Social disadvantage index utilization greatly expanded during the COVID-19 pandemic given the need for tools to rapidly facilitate equitable vaccine allocation. In spring 2021, a presidential executive order directed federal agencies to use these indices to address inequities in vaccine programs,^8^ and most states utilized at least one index to inform COVID-19 vaccine distribution strategies.^9^

Variables in social disadvantage indices are typically obtained from publicly available sources, such as the American Community Survey (ACS).^10^ For example, the Center for Disease Control’s Social Vulnerability Index (SVI) and the Area Deprivation Index (ADI) are two frequently utilized indices in health policy and health services research. These indices were developed to guide resource allocation during disasters (SVI) and inform health care delivery and policy (ADI), respectively.^11,12^ However, indices are often used interchangeably or based on ease of access or geographic level rather than the question at hand or data included. The Child Opportunity Index (COI) was developed using a conceptual model of child development and contains variables such as educational outcomes and access to healthy food.^13^ The COI may be a more suitable metric for questions about child health, but it is unknown whether the COI performs differently than other indices in practice.^14^

While associations between social disadvantage and health outcomes are often described,^15–19^ it is unclear whether associations vary depending on choice of social disadvantage index. While some research has compared the utilization of various indices in the context of COVID-19,^20^ results have been mixed. Our objective was to compare associations across multiple social disadvantage indices and child health outcomes to better inform the choice of social disadvantage index in pediatric research and public health practice. We included three disadvantage indices—the Area Deprivation Index (ADI), the Social Vulnerability Index (SVI), and the Child Opportunity Index (COI). We compared associations between these indices and two child health outcomes impacted by social disadvantage through different mechanisms— infant well-child check (WCC) attendance and adolescent obesity. Because the COI is the only index designed to quantify the community-level vulnerability experienced by children, we hypothesized that associations between the COI and child health outcomes would differ from associations between the ADI or SVI and the same outcomes.

## Methods

### Study Population

We used Electronic Health Record (EHR) data from Duke University Health System (DUHS) from January 1, 2014 to December 31, 2019. DUHS is a comprehensive medical system consisting of three hospitals and a network of primary and specialty care clinics. The source population included patients ≤18 years old, with outpatient encounters between January 1, 2014 and December 31, 2019, and who were Durham County residents (Figure 1). Durham County is the sixth-largest county in North Carolina (NC), with a population of approximately 320,000.^21^ In 2018, the overall median household income in Durham County was $76,962, but $44,004 for Hispanic households and $42,417 for Black households.^22^ Child poverty is higher than adult poverty in Durham County, with almost one fifth of children living in poverty.

**Figure 1:**
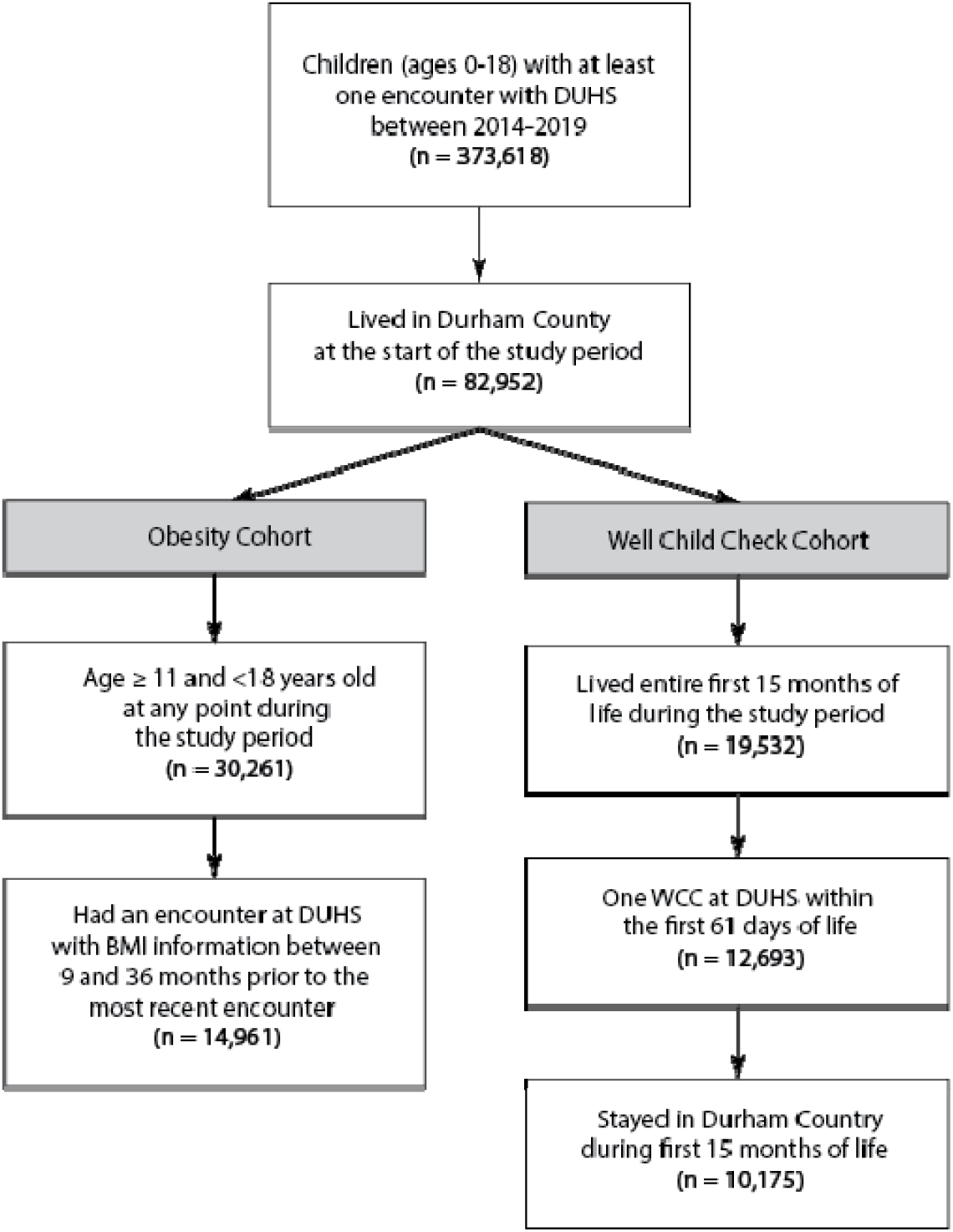
Inclusion Criteria For Patients In WCC And Obesity Cohorts.

**Figure 2.**
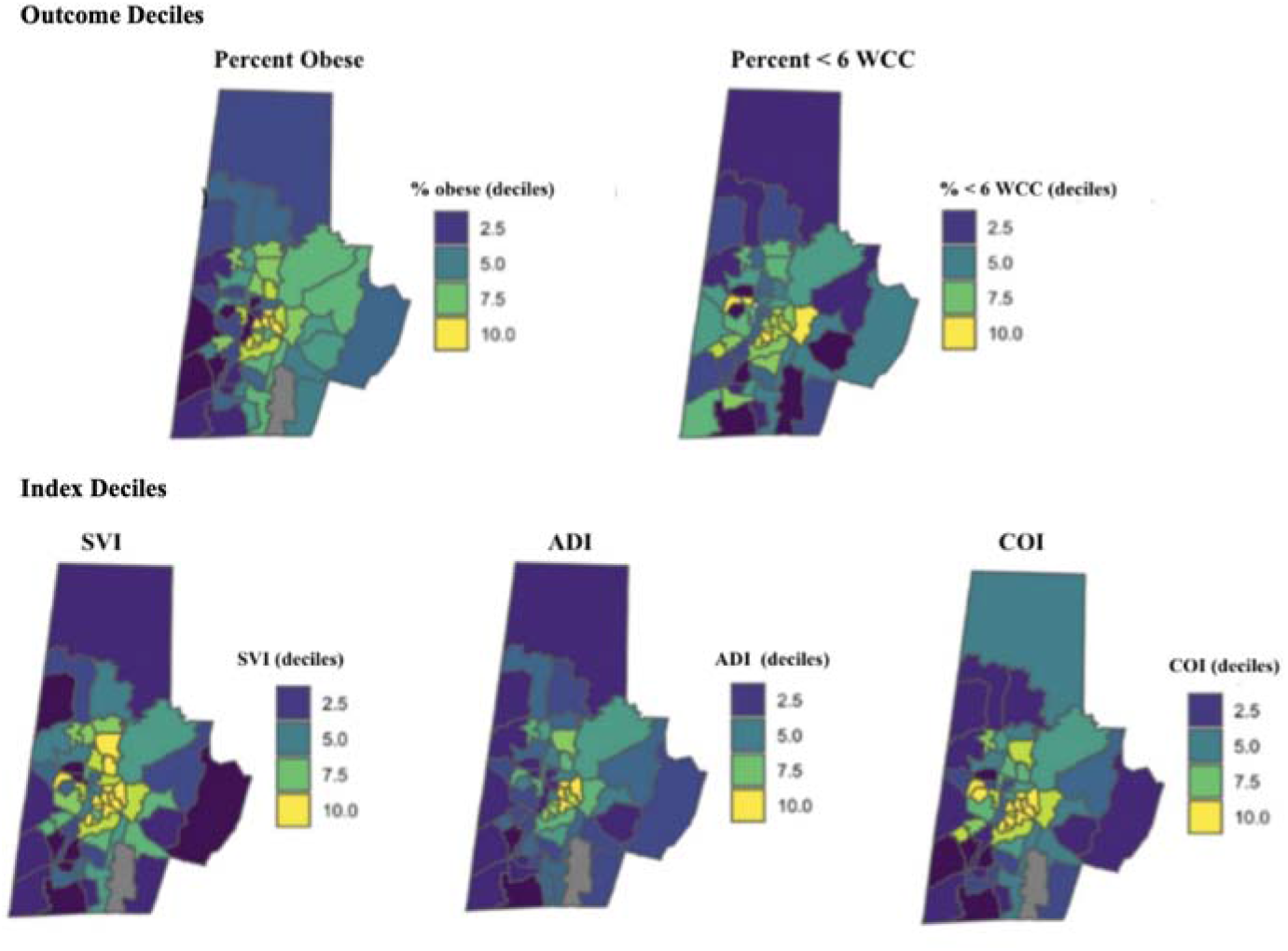
Comparing Obesity & WCC Deciles to Indices Deciles at Census Tract Level. In top row of maps, darker colors represent higher outcome prevalence. In bottom row, darker colors indicate greater disadvantage. ADI rolled up to census tract level to align with COI and SVI geographies; COI recoded so that so that higher scores indicate greater disadvantage as with the other indices.

Patients were required to have non-missing address information. All encounters for patients who did not live in Durham County at the beginning of the study (1/01/2014) were excluded. For patients who moved out of Durham County or between census tracts during the study period, encounters after the first move were excluded. Separate cohorts were developed for obesity and WCC outcomes.

### Exposures

Three disadvantage indices were included: the 2014 SVI, the 2015 ADI, and the 2015 COI 2.0 (Table 1). Years were chosen based on index availability and so that that index values reflected social disadvantage during the same time frames to the closest extent possible. The SVI is available at the census tract level and ranks each tract using 15 social factors obtained from the ACS ^11^ These factors are grouped into four themes: socioeconomic status, household composition and disability, minority status and language, and housing type and transportation. The ADI, which also uses data from the ACS, is available at the census block group level and includes 17 indicators in domains of employment, income, education, and housing quality.^12^ The Child Opportunity Index 2.0, which is available at the census tract level, is based on 29 indicators spanning three domains: education, health and environment, and social and economic.^13^ Unlike the SVI and ADI, the COI draws from multiple data sources, some public and some proprietary, including the ACS, the National Center for Health Statistics, the Environmental Protection Agency, and two proprietary educational datasets.

**Table 1.** Components of Each Vulnerability Index.

| SVI | ADI | COI |
| --- | --- | --- |
| **Socioeconomic Status**<br>Families below the poverty line<br>Unemployment rate<br>Per capita income<br>No high school diploma | **Education**<br>Population aged $\geq 25$ years with $< 9$ years of education<br>Population aged $\geq 25$ years with $<$ a high school diploma<br>Employed persons $\geq 16$ years of age in white-collar occupations | **Early Childhood Education**<br>ECE centers<br>High quality centers<br>ECE enrollment |
| **Household composition & disability**<br>Aged 65 or older<br>Aged 17 or younger<br>Older than age 5 with a disability<br>Single-parent households | **Income & Employment**<br>Median family income<br>Income disparity<br>Unemployment rate<br>Families below the poverty line<br>Population below 150% of the poverty threshold | **Elementary Education**<br>Third grade reading<br>Third grade math |
| **Minority status & language**<br>Minority<br>Speak English “less than well” |  | **Secondary and postsecondary Education**<br>Graduation rate<br>AP course enrollment<br>College enrollment |
| **Housing type & transportation**<br>Multi-unit structures<br>Mobile homes<br>Crowding<br>No vehicle<br>Group quarters | **Housing**<br>Median home value<br>Median gross rent<br>Median monthly mortgage<br>Owner-occupied housing<br>Housing units without complete plumbing | **Educational and social resources**<br>School poverty<br>Teacher experience<br>Adult ed. attainment |
|  | **Household Characteristics**<br>Single-parent households<br>Vehicle ownership<br>Telephone ownership<br>Plumbing in housing units<br>Crowding | **Healthy environments**<br>Access to healthy food<br>Access to green space<br>Walkability<br>Housing vacancy rate |
|  |  | **Toxic Exposures**<br>Hazardous waste sites<br>Industrial pollutants<br>Airborne microparticles<br>Ozone concentration<br>Extreme heat exposure |
|  |  | **Health resources**<br>Insurance coverage |
|  |  | **Economic opportunities**<br>Unemployment rate<br>Commute duration |
|  |  | **Economic and social resources**<br>Poverty rate<br>Public assistance rate<br>Homeownership rate<br>High-skill employment<br>Median household income<br>Single-headed households |

### Outcomes

#### Infant Well-Child Check (WCC) attendance

Infant WCC attendance is a proxy measure for access to care; patients with greater HRSNs (e.g. lower income, transportation barriers, housing instability) are more likely to miss WCCs.^23^ Children were eligible if they were born between 1/01/2014 and 9/30/2018 and therefore had lived their entire first 15 months of life during the study period (Figure 1). Children were required to have established care, defined as having a WCC at DUHS during the first 61 days of life. The outcome was based on the Healthcare Effectiveness Data and Information Set (HEDIS) pediatric quality measures, which specify that infants should attend at least 6 WCC in the first 15 months of life.^24^ The outcome was coded as a binary variable and quantifies the proportion of children who did not attend ≥ six WCCs during the first 15 months of life. Children who moved during their first 15 months of life were excluded.

#### Adolescent obesity

Adolescent obesity is a chronic condition that may reflect accumulated exposure to disadvantage, and is associated with HRSNs including health insurance coverage and parental educational attainment.^25,26^ The cohort included children aged 11-17 years old during the study period. For children who aged into or out of the cohort, only encounters within the 11-17-year range were included. Encounters with missing BMI information were excluded. Obesity was defined as having a BMI ≥ the 95th percentile at the patient’s most recent encounter with non-missing BMI information and at a second encounter ≥ 9 months before and within 36 months of that encounter.^27^

#### Statistical Analysis

Patient addresses were linked to the 2014 SVI, the 2015 ADI, and the 2015 COI 2.0 using Federal Information Processes (FIPs) codes. We used a state-normed version of each index, meaning that percentiles assigned to each geographic area were based on the ranking of that area compared to all others in NC. The output of the SVI is a percentile score from 0-1; a score closer to 1 indicates greater disadvantage. The output of the COI is a score from 1-100, where 1 indicates low opportunity or high disadvantage and 100 indicates high opportunity or low disadvantage. We aggregated SVI and COI scores into deciles and reverse-coded the COI so that a higher score indicated higher disadvantage. The state-normed ADI assigns a score from 1-10, with 1 corresponding to the lowest level of disadvantage and 10 indicating the highest level of disadvantage, to each census block group (subdivision of census tracts). We used a population-weighted mean function to assign ADI scores to census tracts.^28^

Multivariable logistic regression models were used to quantify the associations between each index and outcome. Models were adjusted for race, ethnicity, and sex. Age was included as a covariate for the obesity model, but not for the WCC model because age is included in the WCC outcome definition. Age was defined at the time of the most recent encounter with non-missing BMI information. All analysis was conducted in R.^29^

## Results

The dataset contained 373618 patients with 3816345 encounters at DUHS from January 1, 2014 to December 31, 2019 (Figure 1). Of these, 82952 patients started the study period living in Durham County, and these patients had 1022437 encounters at DUHS, excluding encounters after patients moved. The median (IQR) decile SVI, ADI, and COI for Durham County tracts were 5 (2-9), 4 (2-6.75), and 5 (2-9), respectively.

### WCC Attendance

A total of 10175 patients were included in the WCC cohort. Of these, 4966 (51.2%) were female, 3712 (36.5%) were White, 3363 (33%) were Black, and 477 (4.7%) were Asian. There were 1714 (16.8%) Hispanic/Latino patients (Table 2).

**Table 2:**
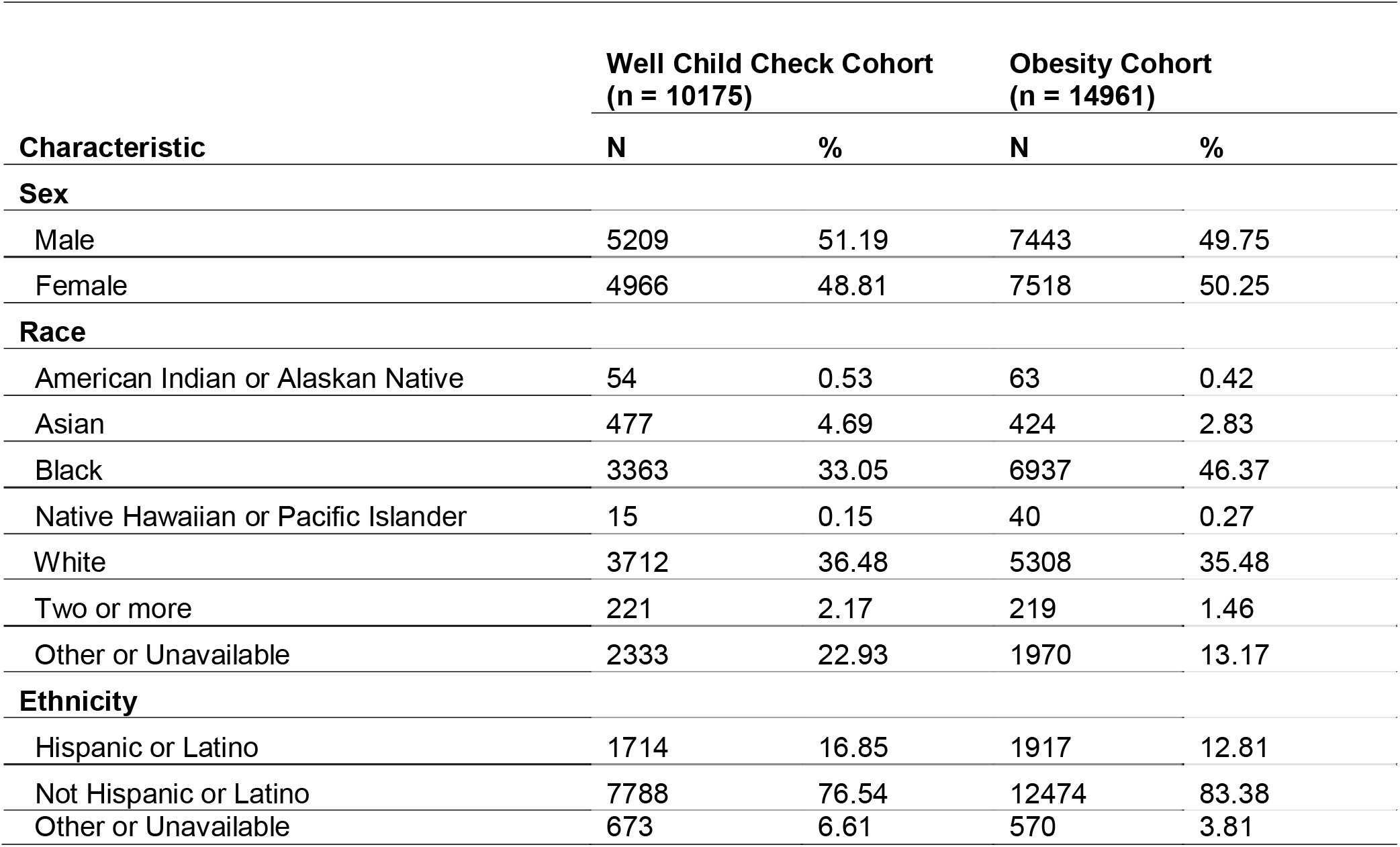
Sample Characteristics Among Patients in WCC And Obesity Cohorts.

| **Characteristic** | **Well Child Check Cohort**<br>**(n = 10175)** |  | **Obesity Cohort**<br>**(n = 14961)** |  |
| --- | --- | --- | --- | --- |
|  | **N** | **%** | **N** | **%** |
| **Sex** |  |  |  |  |
| Male | 5209 | 51.19 | 7443 | 49.75 |
| Female | 4966 | 48.81 | 7518 | 50.25 |
| **Race** |  |  |  |  |
| American Indian or Alaskan Native | 54 | 0.53 | 63 | 0.42 |
| Asian | 477 | 4.69 | 424 | 2.83 |
| Black | 3363 | 33.05 | 6937 | 46.37 |
| Native Hawaiian or Pacific Islander | 15 | 0.15 | 40 | 0.27 |
| White | 3712 | 36.48 | 5308 | 35.48 |
| Two or more | 221 | 2.17 | 219 | 1.46 |
| Other or Unavailable | 2333 | 22.93 | 1970 | 13.17 |
| **Ethnicity** |  |  |  |  |
| Hispanic or Latino | 1714 | 16.85 | 1917 | 12.81 |
| Not Hispanic or Latino | 7788 | 76.54 | 12474 | 83.38 |
| Other or Unavailable | 673 | 6.61 | 570 | 3.81 |

Of the 10175 patients in the WCC cohort, 20% (n = 2073) had less than six WCCs in the first 15 months of life. After adjusting for race, sex, and ethnicity, a one decile increase in SVI was associated with 10% greater odds of having fewer than six WCCs (OR 1.10; 95% CI 1.08-1.12) (Table 3). A one decile increase in ADI was also associated with 10% greater odds of having fewer than six WCCs (OR 1.10; 95% CI 1.08-1.12). A one decile increase in COI was associated with 12% greater odds of having less than six WCCs (OR 1.12; 95% CI 1.10-1.14). All associations were significant (p < 0.001).

**Table 3:** Adjusted Odds of < 6 Well Child Checks in First 15 Months of Life or Obesity by Vulnerability Index.

| **Index** | **Well Child Check Measure <sup>a</sup>** | **Obese** |
| --- | --- | --- |
|  | **Adjusted OR (95% CI)** | **Adjusted OR (95% CI)** |
| SVI | 1.10 (1.08-1.12) | 1.05 (1.04-1.08) |
| ADI | 1.10 (1.08-1.12) | 1.08 (1.06-1.10) |
| COI | 1.12 (1.10-1.14) | 1.07 (1.05-1.08) |

ADI rolled up to census tract level to align with COI and SVI geographies; COI recoded so that all indices coded so that higher scores indicate greater disadvantage.

### Obesity

A total of 14961 patients were included in the obesity cohort. Of these, 7443 (49.7%) were female, 6937 (46.4%) were Black, 5308 (35.5%) were white, and 424 (2.83%) were Asian; 1917 (12.8%) patients were Hispanic or Latino (Table 2).

Of the 14961 patients in the obesity cohort, 20% (n = 2933) qualified as obese. After adjusting for age, race, sex, and ethnicity, a one decile increase in SVI was associated with 5% greater odds of obesity (OR, 1.05; 95% CI, 1.04-1.08) (Table 3). A one decile increase in ADI was associated with 8% greater odds of obesity (OR, 1.08; 95% CI 1.06-1.10); a one decile increase in COI was associated with 7% greater odds of obesity (OR, 1.07; 95% CI 1.05-1.08). All associations were significant (p < 0.001).

### Comparison Across Indices

The association between each index (SVI, ADI, and COI) and each outcome (WCC attendance and adolescent obesity) was statistically significant (Table 3). A one-decile increase in each index was associated with 10-12% greater odds of attending fewer than six infant WCC; each one-decile increase was associated with 5-8% greater odds of adolescent obesity (Figure 2). For both obesity and WCCs, the confidence intervals for all three associations overlapped.

Subgroup analyses were conducted in which the population was stratified by race, sex, and ethnicity to investigate whether index-outcome associations varied by population group. No appreciable differences were detected between results of the stratified analyses and main analysis. Sensitivity analyses in which social disadvantage indices were coded as binary measures (i.e. index score dichotomized at the median score) yielded no relevant differences.

## Discussion

In this study comparing the use of three frequently used social disadvantage indices, we found similar associations across all three indices and both infant WCC attendance and adolescent obesity. As both outcomes are known to be associated with multiple HRSNs, our findings that each index was associated with these outcomes was expected. However, despite differences in index construction, content, and purpose, the magnitudes of associations were similar across the different indices, including the child-specific COI. Our results suggest that—at least in some situations—these three social disadvantage indices may be comparable composite summaries of neighborhood deprivation.

Our findings contribute to an emerging body of mixed evidence assessing the associations between multiple social disadvantage indices and one or more health outcomes. In a study comparing associations between the ADI, SVI, and two other social disadvantage indices (the COVID-19 Community Vulnerability Index and the Minority Health-Social Vulnerability Index) with COVID-19 outcomes in the United States, the ADI had a stronger association with COVID-19 mortality than the other indices.^20^ One large index-outcome comparison study used publicly available county-level data to compare five social disadvantage indices to 24 health outcomes. The ADI and COI were most strongly associated with the greatest number of outcomes and authors concluded that the SVI performed moderately worse compared to the other two indices.^30^ In contrast, the COI and SVI were similarly associated with early and middle childhood obesity in a large national cohort, similar to the results of the present study.^16^ The variation between these findings may be a result of differing geographic units (e.g. county-level versus census tract), the use of individual-level variables in some studies, or differences in population characteristics. Factors such as the particular outcome of interest or population under study are also likely to impact the comparability of disadvantage indices. It may be helpful for index utilizers to consider the intended purpose of a given index and assess whether the variables included are relevant for a particular research question or policy decision. The ADI was originally created for use in health and health outcomes research, while the SVI was created by the CDC to inform disaster response, and the COI incorporates factors that specifically impact children. The variables in each index, and in some cases the weights assigned to each variable, are related to their intended use cases. Users might also consider potential tradeoffs around data accessibility—the ADI and SVI are based on publicly available census data and updated regularly, whereas the COI includes less easily obtainable proprietary information which is updated less consistently.

Similarities in the variables used to create the three disadvantage indices included in this study may explain our findings. All three indices share four variables representing poverty rate, high school education attainment, single-headed households, and unemployment. All three also include a measure of income, although ADI and COI use median household income, while the SVI includes per capita income. Although several variables, such as measures of crowding and vehicle ownership, are shared between the SVI and the ADI, the COI contains unique variables related to education, food access and environmental exposures. Despite these differences, variables shared between the indices may be closely correlated with variables unique to each index, so that unique variables did not make a significant difference in the associations examined here. For example, measures of income are known to be closely related to educational resources available in many communities.^31,32^ It is plausible that the income variables present in all three indices and the unique child-specific education variables included in the COI similarly contribute to the associations between social disadvantage and the health outcomes considered here.

Our findings suggest considerations for researchers or policymakers utilizing social disadvantage indices. Given the multidimensionality of the relationship between neighborhood deprivation and health, understanding the causal pathways influencing specific health outcomes can help guide index choice. While the COI performed comparably to the ADI and SVI here, associations with outcomes more closely linked to environmental exposures (e.g. respiratory health) than infant WCC attendance or adolescent obesity may be less biased with the COI. Geographic differences in social policies may also impact the way social disadvantage indices are associated with health outcomes. For example, in our study, income and education variables might be highly correlated. However, this relationship may vary outside of NC depending on education policies—18 states have social policies to correct for inequity between high-and-low-poverty school districts due to differences in property tax-derived funding.^33^ Last, although the indices considered here are frequently utilized to inform health equity initiatives, these indices were not designed to measure the impacts of structural racism. Of the indices utilized in this study, only the SVI includes an indicator of neighborhood racial and ethnic composition. Emerging research suggests that indices like the ADI which do not incorporate race or ethnicity may still capture the impacts of structural racism on health through inclusion of variables reflecting residential segregation, such as access to parks or retail availability.^34^ However, a simulation study showed that using the SVI as opposed to the ADI for COVID-19 vaccine allocation resulted in more resources being distributed to minority populations.^35^ Future research should explore the potential implications of excluding measures of racial or ethnic composition when choosing between indices. For studies or programs targeting racial health inequities, explicit measures of structural racism should be considered alongside measures of neighborhood disadvantage.^4,36^

## Strengths and Limitations

This study has some limitations. We included a cohort of children from a single county, so external validity may be limited. While the Durham County population is diverse as measured by race and ethnicity, wealth, and educational attainment, results may differ in more homogenous populations. The use of EHR data, and our inclusion criteria that required establishing care in the WCC attendance cohort, and multiple visits in the obesity outcome cohort, means that the study population reflects patients with health care engagement. Finally, we did not account for the effects of geographic mobility on the relationship between social disadvantage and health outcomes.

Our study has notable strengths. We included a large sample of diverse children in each cohort, which was facilitated by use of EHR data. Additionally, because we selected three commonly used social disadvantage indices that have been previously used in both pediatric and adult populations, findings will be broadly applicable to both researchers and policymakers.

## Conclusions

We found similar associations between increased social disadvantage, as defined by three different indices, and both decreased WCC attendance and adolescent obesity. The magnitude of association between the health outcomes and the child-centered COI did not differ from other indices. While the COI includes variables specific to children, it may be most useful in quantifying associations driven by factors not incorporated into other indices. All three indices can inform policy and practice strategies that promote child health equity. However, given mixed evidence around how choice of index may influence associations, public health leaders and researchers should continue to critically assess the content included, pathways influencing outcomes of interest, and data availability when using social disadvantage indices.

## Data Availability

Analytic data or code can be made available upon reasonable request to the authors.

## Article Information

## Author Contributions

Ms. Zolotor had full access to all of the data in the study and takes responsibility for the integrity of the data and the accuracy of the data analysis. Drs Bhavsar and Cholera shared last authorship.

### Concept and design

Zolotor, Cholera, Bhavsar. *Acquisition, analysis, or interpretation of data*: All authors.

### Drafting of the manuscript

Zolotor, Cholera

### Critical revision of the manuscript for important intellectual content

All authors.

### Statistical analysis

Zolotor, Huang.

### Administrative, technical, or material support

Cholera, Bhavsar

### Supervision

Cholera

## Data Sharing Statement

Data or analytic code can be made available upon reasonable request to the authors.

## Additional contributions

We thank Congwen Zhao, MS, a biostatistician at the Duke University School of Medicine, for her support with data acquisition and queries. We also thank Kamaria Kaalund, BA; Greeshma James, MPH; Charles Wood, MD MPH; and Reilly Dever, MD MBA for their contributions to the study concept and design.

